# Cost-effectiveness of vector control strategies for supplementing mass drug administration for eliminating lymphatic filariasis in India

**DOI:** 10.1101/2023.12.10.23299653

**Authors:** Donald S. Shepard, Aung K. Lwin, Sunish I. Pulikkottil, Mariapillai Kalimuthu, Natarajan Arunachalam, Brij K. Tyagi, Graham B. White

## Abstract

**Background/Methodology:** Despite progress using mass drug administration (MDA), lymphatic filariasis (LF) remains a major public health issue in India. Vector control (VC) is hypothesized as a potentially useful addition to MDA towards LF elimination. We conducted cost-effectiveness analysis of MDA alone and augmented by VC single (VCS) or integrated VC approaches (VCI). Data came from historical controls and a 3-arm cluster randomized trial of 36 villages at risk of LF transmission in Tamil Nadu, India. The arms were: MDA alone (the standard of care); VCS (MDA plus expanded polystyrene beads for covering the water surface in wells and cesspits to suppress the filariasis vector mosquito *Culex quinquefasciatus*), and VCI (VCS plus insecticidal pyrethroid impregnated curtains over windows, doors, and eaves). Economic costs in 2010 US$ combined government and community inputs from household to state levels. Outcomes were controlled microfilaria prevalence (MfP) and antigen prevalence (AgP) to conventional elimination targets (MfP<1% and AgP<2%) from 2010 to 2013, and disability adjusted life years (DALYs) averted.

**Principal Findings:** The estimated annual economic cost per resident was US$0.53 for MDA alone, US$1.02 for VCS, and US$1.83 for VCI. With MDA offered in all arms, all reduced LF prevalence substantially and significantly from 2010 to 2013. MDA proved highly cost effective at $112 per DALY, a very small (8%) share of India’s then per capita GDP. Progress towards elimination was comparable across all three study arms.

**Conclusions:** The well-functioning MDA was effective and very cost-effective for eliminating LF, leaving little scope for further improvement. Supplementary VC demonstrated no statistically significant additional benefit in this trial.

**Authors’ Summary:** Lymphatic filariasis (LF) is one of the twenty neglected tropical diseases (NTDs) that affect more than one billion people worldwide. As part of the effort toward global elimination of LF, the Indian State of Tamil Nadu government has implemented mass drug administration (MDA) repeatedly since 1996. Despite their efforts, LF had not yet been eliminated. Although vector control (VC) is proposed to augment regular MDA to help eliminate LF, little is known about the increased impact or costs. Our study compares the costs of MDA alone to the combination of MDA with alternative VC interventions. We calculated both program operating costs and costs to communities. We found MDA to be very cost-effective for eliminating LF. Against low levels of LF endemicity (≤10%), the study had limited ability to detect further improvements and found no significant incremental improvements from VC.

## Introduction

Among the neglected tropical diseases [1] prevalent in India, lymphatic filariasis (LF) remains a major public health problem [2–5]. This chronic disease is characterized by acute episodes of fever with painful lymph nodes (adenolymphangitis) and progressive swelling of lower and/or upper limbs (lymphoedema) and testes (hydrocele), resulting in loss of labor and depleting the quality of life for those infected [6]. The latest data show substantial reductions in LF prevalence in most endemic countries and LF elimination from 20 countries [7], but still the persistence of a major global problem [8]. To prevent this debilitating disease in India, the country launched the National Filaria Control Programme in 1955 [9]. Control of LF is based on mass drug administration (MDA), where local workers give an annual dose of preventive medicines to all residents of the target area.

The Indian State of Tamil Nadu, with a strong public health infrastructure, began MDA in 1996 and gradually extended the program to cover all LF endemic districts [4]. Initially the MDA program administered a single dose of diethylcarbamazine (DEC) to each eligible person; starting in 2000, the program administered DEC plus albendazole annually to the whole population of districts (implementation units) where LF was endemic [10, 11]. In 2015, MDA covered 255 districts of 21 states and territories in India where 630 million people are at risk of LF transmission [2]. However, even after 5 rounds of MDA with both drugs, pockets of LF infection persist [12], attributed to inadequate MDA coverage, likely due to non-compliance with timely ingestion of the free antifilarial drugs [13, 14].

LF is mostly caused by the nematode worm *Wuchereria bancrofti*, transmitted in India by vector *Culex quinquefasciatus* mosquitoes [15, 16] which develop in polluted water, such as water drained into cesspits and sewers [17]. Implementing vector control (VC) programs to interrupt filariasis transmission is considered a potentially beneficial supplement to MDA programs [18, 19], perhaps essential towards the elimination of LF [20, 21]. Epidemiological evidence of the benefit from supplementary VC [22, 23] reported that two annual rounds of MDA augmented by VC with expanded polystyrene beads (EPB) and larvivorous fishes to control *Culex* aquatic stages reduced both the annual transmission potential and the transmission intensity index of LF to zero in Tamil Nadu villages [24]. A cost-effectiveness study determined that adding VC with two annual rounds of MDA decreased the prevalence of microfilaremia by 1% with an added cost of US$0.85 per person annually [25].

In 2009, the Indian Council for Medical Research Centre for Research in Medical Entomology (CRME) commenced this prospective randomized trial in Tamil Nadu to examine the effects of specific VC interventions for augmenting MDA, compared with the regular MDA program alone. Both VC and MDA strengthened social cohesion by combining community-based initiatives, multiple levels of Indian government, and global partners. The results showed that all of the interventions were associated with significant reductions in LF [26, 27], highlighting the need for further analysis to clarify the best strategy. As LF has persisted both globally and specifically in India, the global health community, donors, and national governments have continued searching for affordable interventions to eliminate LF. Emerging problems, such as controlling the COVID-19 pandemic, compete for global public health resources. Cost constraints affect all levels--local communities, national governments, and global institutions. LF, one of the most neglected diseases, and COVID-19, currently one of the most salient global problems, both illustrate the challenges of implementing technologies with available resources.

Despite decades of effort, LF continues to affect the most marginalized communities. It is not clear whether the existing strategy, MDA, will be sufficient to eliminate LF as a public health problem. To our knowledge, Sunish et al. [26, 27] conducted the only controlled study of effectiveness of augmenting MDA with VC. Sound policies depend not only on effectiveness, but also on costs. To determine the best use of available resources, this cost-effectiveness study assessed the costs of the comparative interventions of Sunish et al.[26, 27] and built on previous research by CRME [22, 23, 28]. Our study measures the costs and effectiveness of MDA, assesses the incremental costs of VC interventions, and computes the incremental cost-effectiveness ratio of adding VC interventions compared to regular MDA alone. With its rigorous assessment of costs and outcomes, we feel that this study offers key information about future LF control.

## Methods

### Setting

The State of Tamil Nadu had a population of 72 million persons in 2011 [29]. Lymphatic filariasis was endemic in at least 20 of its 33 districts [30]. The State Health Department funds health services in all districts. In 2008-09, the public system contained 42 health unit districts (HUDs) which supervised 1,421 primary health care (PHC) units in Tamil Nadu, with each PHC serving about 30,000 residents [31]. This cluster randomized trial collected data from 2010 to 2013 in Kallakurichi Health Unit District [32], which had a population of 3.5 million residents and 36 PHC units. The CRME, with headquarters in Madurai, Tamil Nadu, opened a field station in the Tirukoilur block of the Kallakurichi Health Unit of Villupurum [22]. The entomological team selected 36 villages as study sites based on their high risk of LF (Figure 1). With a combined population of 43,000 people, they were under three PHC units in the Tirukoilur block: Ariyur, Edaiyur and T.Kunnathur. Altogether, these three PHCs provided health care for about 150,000 people.

**Figure 1:**
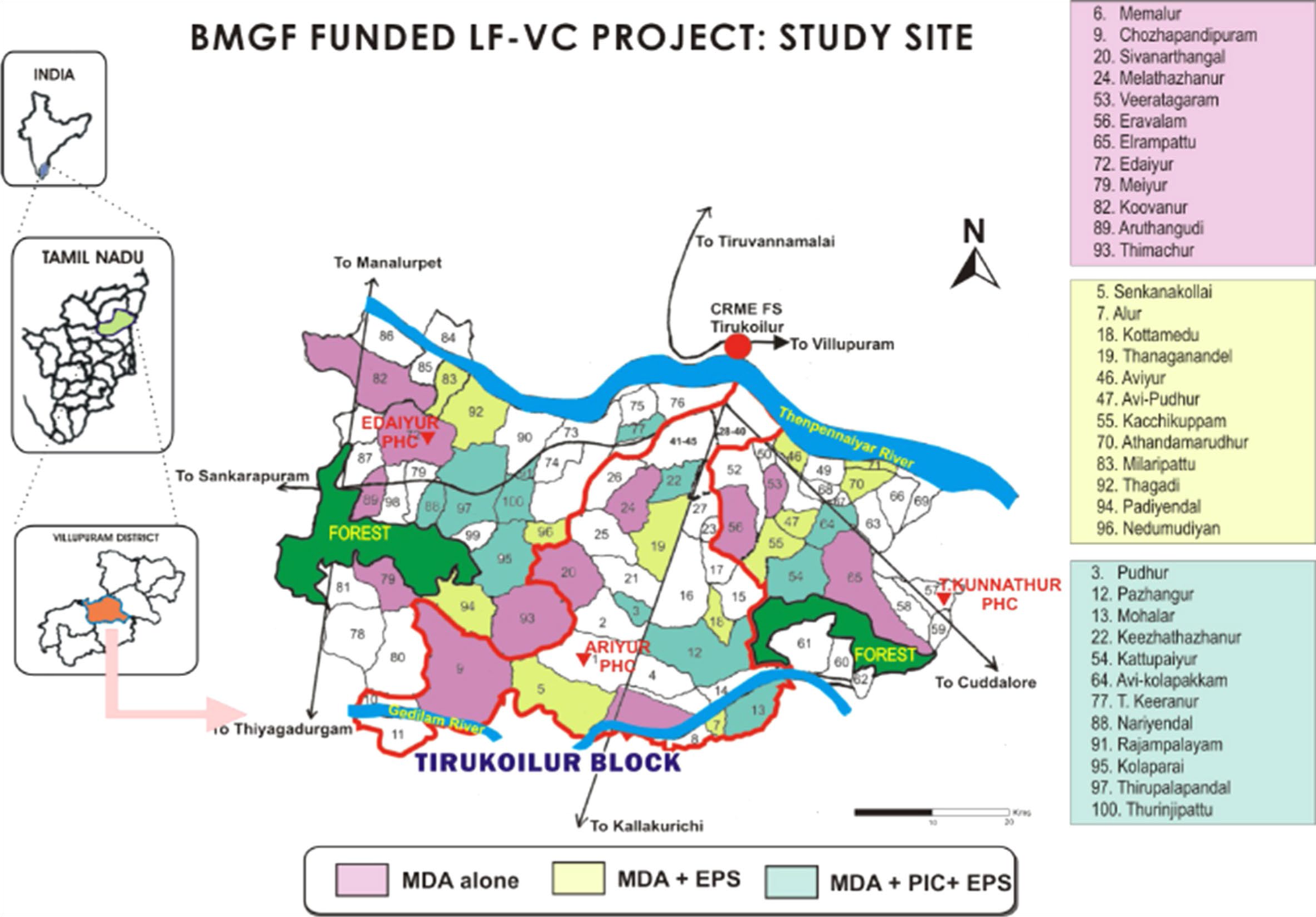
Map of study villages by treatment arm in lymphatic filariasis vector control (LF-VC) project funded by the Bill & Melinda Gates Foundation (BMGF)

The CRME randomly assigned 12 villages to each of the three study conditions. The allocation was balanced exactly by LF microfilaremia baseline prevalence (MfP) and ecological circumstances for mosquitoes: 3 villages with MfP<1%, 3 villages with MfP 1-2%, and 6 villages with MfP>2%; and agreed closely in distribution by condition within each PHC block. The World Health Organization [33] defined the eligibility of communities for MDA treatment as the prevalence of filarial parasites (MfP) or antigen of more than 1%. While the population of villages varied [mean (M) = 1914, standard deviation (SD) =897], all villages included in this study were eligible for treatment (M = 85%, SD = 9%) with the WHO’s recommended two-drug regimen of DEC and albendazole [34].

Kallakurichi Health Unit District lies in northern Tamil Nadu, 40 to 80 kilometers inland from Puducherry on the Indian east coast. The general climate in the region is hot and dry, with moderate rainfall from September to December [35]. The majority of villagers are landless laborers who earn their living from agriculture and livestock [28]. For community management and administration, village committees are led by elected village presidents. Government health personnel contact with villagers relies on the village health nurse, who manages a PHC satellite unit called a sub-center, that serves approximately 5,000 inhabitants in two to three villages [36].

Prior to the study, no systematic VC operations had been routinely implemented in the study area. Villages have abandoned wells, since tap water standpipes have been installed. Sanitation involves pit latrines. The majority of houses lack effective barriers to insects, allowing endophilic mosquitoes easy access into the interior of dwellings and thus are able to bite residents inside. Due to the region’s hot and humid weather, villagers mostly prefer not to use bed nets for the prevention of mosquito bites, because they reduce ventilation. Traditionally, water used for showering and cleaning utensils is disposed of in soakaway pits and wet ditches, which readily serve as breeding sites for *Culex* mosquitoes.

### Study design

The State Health Department (SHD) of Tamil Nadu implements MDA almost annually with tablets of DEC, procured from local manufacturers in India [4], plus albendazole donated by the pharmaceutical company GlaxoSmithKline [37], following procedures and dosage regimens recommended by the Global Program to Eliminate Lymphatic Filariasis [38]. In the study area, the SHD organized the local program to eliminate lymphatic filariasis through five management levels: the Tamil Nadu SHD, Kallakurichi Health Unit District, primary health centers (PHC), sub-centers, and village delivery units of filariasis prevention assistants.

Drugs for MDA were transported by the SHD to its HUDs, then distributed to their block level PHCs. Village health nurses of sub-centers carried drugs from the block level PHCs to their villages and supervised the most peripheral workers of the delivery chain, the filariasis prevention assistants, who were volunteers. Recruited from village communities, filariasis prevention assistants directly delivered drugs to the households and observed residents swallowing their tablets. In each annual MDA campaign, one day is scheduled for filariasis prevention assistants to deliver to households, and two days are scheduled for tracing and providing treatment of those missed on the preceding day.

Multiple levels of management are required to coordinate delivery of the MDA tablets to residents in the 36 villages investigated. Upper health management levels were involved in more extensive distribution efforts to villages across the State that were not part of this study’s population, although the cost shares of higher-level centers were pro-rated based on their total population coverages. Table 1 shows how a village is integrated into the combined State efforts of all health management levels and associated responsibilities.

**Table 1:** Management levels supporting the LF mass drug administration (MDA) program in the study area and corresponding populations covered.

| Management level | Staff | Population covered |
| --- | --- | --- |
| 1. State Health Department (SHD) of Tamil Nadu | Members of Task Force and Advisory Committees; Supervisory Team | 31,250,000 |
| 2. Health Unit District (HUD) of Kallakurichi | HUD supervisory team: Deputy Director of Health Services, District Staff Assistant, District Malaria Officer, District Health Inspectors*, Medical College Entomologists*, HUD Rapid Response Team; | 1,589,500 |
| 3. Block Primary Health Center (PHC) of Ariyur | Block PHC supervisory team: Block Medical Officer*, Assistant Medical Officer at block, Assistant Medical Officer of additional PHC, Block Health Supervisor, Non-medical Supervisor, Block Extension Educator, Community Health Nurse at block PHC, Sector Health Nurse at block PHC, Health Inspector (MDA in-charge) at Block Health Supervisor*, Non-medical Supervisor , Block Extension Educator*, Community Health Nurse, Sector Health Nurse, Pharmacist*, Sanitary Worker, Driver; Block PHC Rapid Response Team | 116,822 |
| 4. Subcenter | Village Health Nurse * | 5,000 |
| 5. Village | Involved villagers | 1,639 |
| 6. 50 households | Village volunteers * | 250 |

\* Indicates survey respondents. Note: management levels are ordered from highest to lowest. LF denotes lymphatic filariasis.

The VC intervention delivery system for this CRME research study was less hierarchical than that of the National MDA program administered by the state. Major activities of added VC interventions were carried out solely by the CRME team, in close collaboration with Ariyur block PHC staff members, and other field-level government staff for some part of the monitoring and evaluation efforts. CRME recruited volunteers from villages, trained and motivated them for ‘Vector Control through Community’ [27] to facilitate VC activities by community members. While the two higher administrative levels (SHD and HUD) did not perform VC operations during the study, they would manage these activities should the VC programs be adopted nationwide. To make comparisons of interventions more realistic for future purposes, we simulated costs of senior management level efforts in addition to actual efforts of CRME and the contributions from its associated government management centers.

### Description of treatment conditions

The MDA program (intended annually but subject to dates of convenience for administrative and logistical reasons) administered anti-filarial DEC and anthelminthic albendazole to target at-risk communities. DEC, by immune mechanisms still largely undefined, causes rapid clearance and destruction of the filarial larvae (microfilariae) circulating in the blood as well as the slower, progressive death of the adult filarial worms. Albendazole is a broad-spectrum anthelminthic drug that principally affects the adult worms, thereby inhibiting the production of microfilariae and suppressing transmission of LF.

Two interventions were applied for integrated control of the LF vector *Culex quinquefasciatus*. Against the developing stages of *Culex*, a floating layer of expanded polystyrene beads (EPB) was applied at rates of 350–400 g/m² water surface area to the main breeding sites (cesspits and wells) to deter mosquito oviposition and asphyxiate larvae and pupae by keeping them submerged [39]. Against the adult *Culex* mosquitoes, pyrethroid impregnated curtains (PIC) were installed to mosquito landing sites around houses (doors, eaves, and windows) to kill adult insects by contact with insecticidal pyrethroid [40].

Polystyrene resin granules (Shri Sakthi Insulations, Chennai, TN) were boiled to produce the expanded lightweight, ∼2mm diameter EPB [41]. To facilitate the placement and retention of EPB in mosquito breeding sites, it was necessary to modify cesspit frames and unused wells to prevent water overflowing, which would flush away the EPBs. Therefore, cesspits were lined with brick walls, and unused wells were cleaned and covered by a custom-made metal mesh screen in a wooden frame.

The PIC intervention required measurement of the doors, windows and eaves for tailoring curtains to fit these spaces. Curtain materials were made of white woven ventilated polypropylene (VIRGO Polymer India Ltd., Kanchipuram, TN https://virgopolymer.com/fibc/ventilated-fibc) impregnated with deltamethrin insecticide (2.5% SC, Chemet Wets & Flows Pvt. Ltd., Ahmedabad, Gujarat) by soaking and drying in the shade to achieve approximately 25mg ai/m^2^ dosage, before cutting, sewing borders and installation by hanging with nails. Installed curtains had to be re-treated by spraying every six months to sustain insecticidal efficacy (validated by CRME bioassays). CRME implemented both interventions in their assigned villages from March to June 2010.

### Collection of cost data

For both MDA and VC programs, personnel costs were calculated using time allocation surveys, salary and wage records, reports, and narrative interviews with paid and volunteer workers. Reports were used to allocate SHD staff time, while time allocation surveys were completed by managers and some community leaders for all levels below the SHD. The salary of each full-time worker and the opportunity cost of each volunteer were collected from administrative records at the SHD, HUD, three BLPHCs, the CRME headquarters and its field station. Narrative interviews clarified implementation procedures and tasks of health workers and community volunteers. Validated costs of major activities of MDA and VC for each management level of Tamil Nadu’s health system were entered into a customized Excel spreadsheet (Microsoft Corp., Redmond, Washington) cost analysis tool developed to combine the costs of MDA and VC activities from different management levels.

Both MDA and VC involved considerable efforts by community members. Current MDA programs engaged full community mobilization efforts once a year, while VC programs required full community mobilization in the initial year and regular maintenance by villagers in subsequent years. Community mobilization efforts for introducing VC were underway in the study villages at the time of the research team’s visit to India. For a narrative understanding, Brandeis investigators with CRME interpreters interviewed participants at every level from the SHD to households.

Non-personnel items consisted mainly of transportation costs (allocated costs of vehicles and fuel) for all activities, drugs for MDA, polystyrene granules, cooking pots and fire for EPB production, construction materials (bricks, cement, tools) for improving cesspits, insecticide, curtains and tools for cutting and sewing them for PIC. We collected information on non-personnel costs of MDA from administrative records from the PHC, interviews with supervisors, and vector control program reports [42]. To estimate economic costs of donated drugs, we asked informants for local retail prices and validated these against the International Drug Price Indicator Guide, 2009 [43]. For VC interventions, CRME documents and officials provided recorded expenditures on non-personnel items. Authors communicated regularly with CRME Field Unit staff (Supporting information S1 Figure and Supporting information S2 Figure).

### Analysis of cost data

WHO and its international partners such as the United States Agency for International Development (USAID) adopted a 3-stage “roll-out package” [44] against NTDs, assuming that integration and scale-up of various programs to full-national scale is feasible [45]. Our analysis followed similar assumptions that various approaches to eliminating a single NTD can be integrated and scaled up by the national ministries.

In our current LF study, one arm applied only one intervention (regular MDA). The other two arms used combined interventions: VCS combined MDA with EPB and VCI combined MDA, EPB and PIC. We estimated the cost of each intervention separately and assumed that, for a combined program, we could add the components to estimate the final costs of the integrated approaches. Our analysis of unit costs of each intervention consists of three major steps: (1) estimating total recurrent costs (personnel and non-personnel) by management level, (2) annualizing capital costs, and (3) calculating final unit costs per beneficiary.

Microsoft Excel was used to create two cost analysis tools: mass drug administration cost analysis tool (MDA-CAT) and VC cost analysis tool (VC-CAT), which were used to enter the data and perform cost analyses. Raw data from our data collection instruments provided estimates of aggregate costs of personnel activities and non-personnel inputs. We categorized these costs by each management level, i.e., the State Health Department, intermediary management levels, and household groups. As higher levels served populations of varying sizes beyond the study’s 36 villages (Table 1), we converted the costs at each administrative level to their corresponding per capita amounts, including the administrative costs of hiring personnel and procuring materials for VC and MDA. To compare yearly costs across the implementation strategies, we amortized capital assets over their useful lives.

In contrast to the State-run annual MDA programs, the VC activities initiated specifically for this study were managed by CRME and funded with donor support. Community mobilization was most intense in the first year; project managers estimated that only 15% of the staff’s initial year efforts were necessary in subsequent years when communities were presumed to be self-sufficient for continuing their VC efforts. While MDA required the procurement and dissemination of drugs once a year, the lifetime of installed VC materials varied. For example, EPB required yearly production and reapplication, whereas PIC curtains were expected to last for three years, while insecticide spray on curtains needed to be reapplied every six-months.

Our analysis computed per capita costs based on the number of residents benefiting from each service. For MDA interventions, the combination of drugs served one person. For PICs, the curtain benefited one household (average of five persons) while one cesspit modification served 27 households (135 persons). Most cost data on personnel activities and non-personnel inputs were initially collected in Indian rupees. The project was planned, and many costs were incurred in 2010. We therefore converted Indian rupees to US dollars based on the market exchange rate of 45.8 rupees per US dollar in early 2010 [46] and expressed all costs in 2010 US$.

### Analysis of impact

The CRME entomological team collected baseline data of parasitological prevalence and entomological status before the intervention (February 2010) and continued to collect yearly data on the post-intervention progress for three years. We collected data on microfilaria prevalence (MfP) tested by microscopic examination of night-time collection of blood smear samples and AgP using the immunochromatography card test (ICT, BinaxNOW® Filariasis, Alere Inc., Portland, Maine) for MfP and AgP, commonly used indicators to monitor progress towards LF elimination [47–49].

To evaluate the impact of the various control strategies, we first evaluated the impact of the reference strategy, MDA. As MDA is the standard treatment in India, there was on control area without MDA. Instead, we selected as the comparator the historical situation in 2010, as implementation of and compliance with MDA had traditionally been low [50]. We calculated the cumulative improvement from 2010 to 2013 in the share of villages reaching the benchmarks towards elimination of MfP and AgP as public health problems -- MfP<1% and AgP<2%. Dividing by 3 (for 3 years), we then calculated the average annual improvement.

To evaluate the incremental improvement of VC over MDA, we performed logistic regression analyses with these shares of villages controlled based on MfP<1% or AgP<2% as the dependent variable using Stata version 15 (StataCorp, College Station, TX). The independent variables were the final year of intervention (year), the intervention (MDA alone, as the baseline level, VCS, and VCI), and interaction terms. These interaction terms on MfP and AgP (intervention level x year) are the main variables of interest, indicating the differential effectiveness of each combination of interventions after controlling for baseline conditions and general trends. Finally, to contextualize our findings we examined India’s national prevalence of LF from 1990 through 2019, the latest data available.

### Cost-effectiveness analysis

For assessing the cost-effectiveness of VC, we converted the average annual improvements from VC by arm into disability adjusted life years (DALYs) averted per 1,000 population. As the first step, we needed the annual DALY burden per person affected by LF. The one study of which we were aware reported an annual burden per case of 0.11 [51]. However, this relatively high value apparently reflected only symptomatic persons, rather than the broader population of persons infected. To derive an estimate for this broader population, we relied on the Global Burden of Disease (GBD) study. Using the most recent year available (2019), we divided India’s rate of years lost to disability (58.96/100,000) due to LF by its prevalence (2.68%) [52], deriving a burden of 0.022 DALYs per person affected. Finally, we divided the incremental cost per 1,000 population by the incremental DALYs averted per 1,000 population to get the incremental cost-effectiveness ratios (ICER) by arm.

## Results

### Economic cost per person treated

Table 2 summarizes lifespans of the inputs and their target beneficiaries. Table 3 presents the ingredients costs (unit costs and quantities) of each component per 1,000 population at the village level. Table 4 presents the corresponding analysis at a higher level (the block level). Table 5 expands the cost analysis to all levels of the health care system. Table 5 shows that approximately 25% of the economic costs for the MDA program ($110.94 compared to $533.55) were borne by the communities served. From the community perspective, PIC was the costliest intervention, EPB was intermediate, and MDA was least costly. VCI, which combines EPB and PCI, costs $490.93 (i.e., $129.46 plus $361.47) per 1,000 households. This is 4.5 times as much as lowest cost intervention (MDA).

**Table 2:** Useful life spans and targets of MDA and VC inputs.

| Inputs | Useful<br>life<br>(years) | Intervention targets |
| --- | --- | --- |
| <u>Mass drug administration (MDA)</u> |  |  |
| Sensitization by government. | 1 | All residents |
| Drugs | 1 | All residents |
| <u>Vector control (VC)</u> |  |  |
| *Both interventions* |  |  |
| Peer sensitization of villagers | 5 | All residents |
| *Expanded polystyrene beads (EPB)* |  |  |
| Cesspit modification | 3 | Cesspits (1 pit per 27 households) |
| Application of beads | 1 | Wells (1 well per 55 households) |
| *Pyrethroid impregnated curtains (PIC)* |  |  |
| Curtain material | 3 | All houses (1 house per 5 persons) |
| Spray | 0.5 | All houses (1 house per 5 persons) |

**Table 3:**
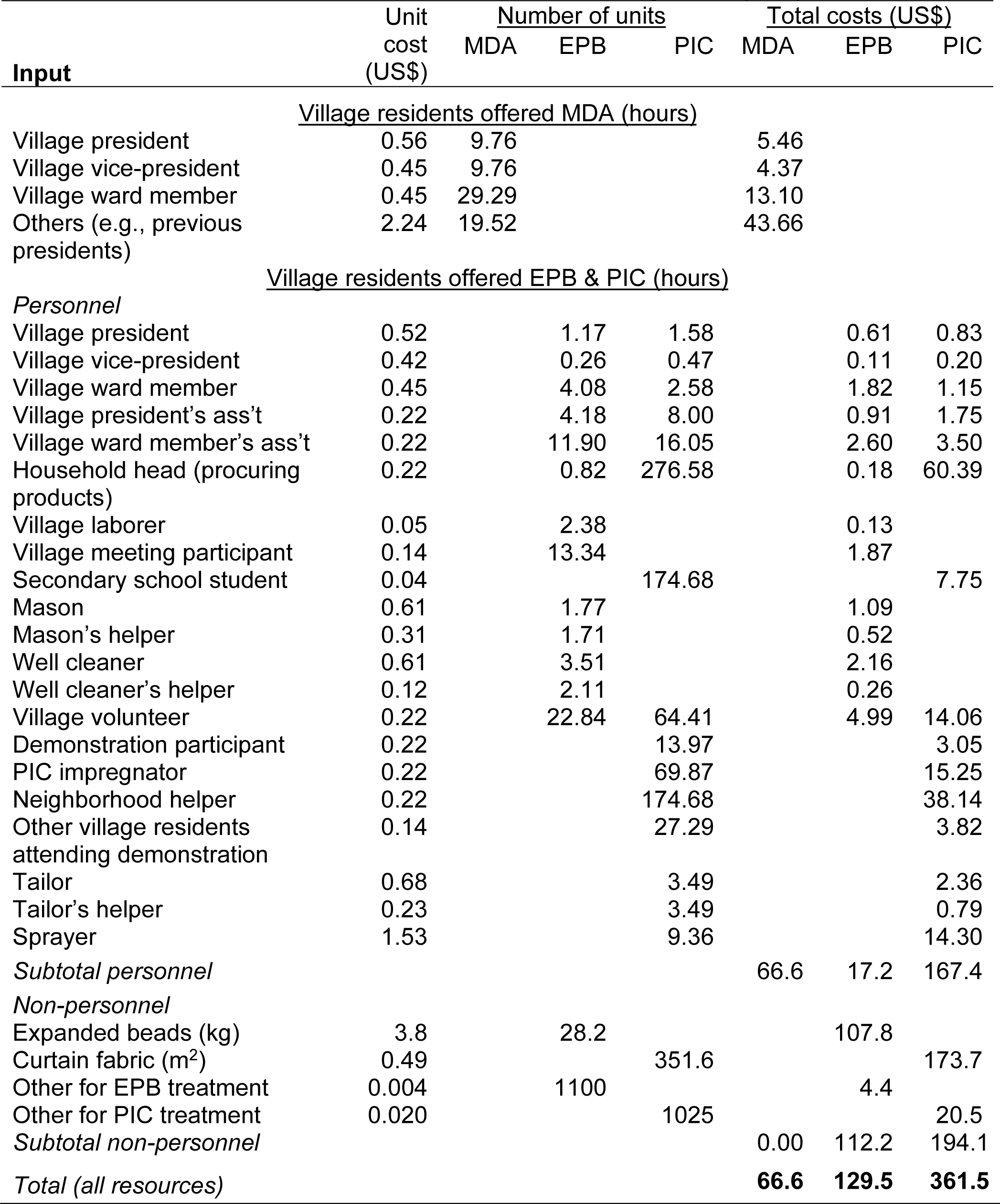
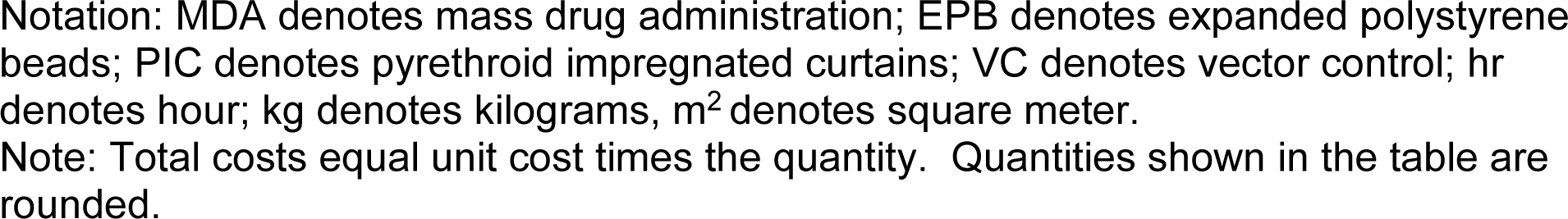
Annualized costs per 1,000 population at the village level (US$)

| Input | Unit<br>cost<br>(US\$) | Number of units | | | Total costs (US\$) | | |
| --- | --- | --- | --- | --- | --- | --- | --- |
|  |  | MDA | EPB | PIC | MDA | EPB | PIC |
| <u>Village residents offered MDA (hours)</u> |  |  |  |  |  |  |  |
| Village president | 0.56 | 9.76 |  |  | 5.46 |  |  |
| Village vice-president | 0.45 | 9.76 |  |  | 4.37 |  |  |
| Village ward member | 0.45 | 29.29 |  |  | 13.10 |  |  |
| Others (e.g., previous presidents) | 2.24 | 19.52 |  |  | 43.66 |  |  |
| <u>Village residents offered EPB & PIC (hours)</u> |  |  |  |  |  |  |  |
| *Personnel* |  |  |  |  |  |  |  |
| Village president | 0.52 |  | 1.17 | 1.58 |  | 0.61 | 0.83 |
| Village vice-president | 0.42 |  | 0.26 | 0.47 |  | 0.11 | 0.20 |
| Village ward member | 0.45 |  | 4.08 | 2.58 |  | 1.82 | 1.15 |
| Village president's ass't | 0.22 |  | 4.18 | 8.00 |  | 0.91 | 1.75 |
| Village ward member's ass't | 0.22 |  | 11.90 | 16.05 |  | 2.60 | 3.50 |
| Household head (procuring products) | 0.22 |  | 0.82 | 276.58 |  | 0.18 | 60.39 |
| Village laborer | 0.05 |  | 2.38 |  |  | 0.13 |  |
| Village meeting participant | 0.14 |  | 13.34 |  |  | 1.87 |  |
| Secondary school student | 0.04 |  |  | 174.68 |  |  | 7.75 |
| Mason | 0.61 |  | 1.77 |  |  | 1.09 |  |
| Mason's helper | 0.31 |  | 1.71 |  |  | 0.52 |  |
| Well cleaner | 0.61 |  | 3.51 |  |  | 2.16 |  |
| Well cleaner's helper | 0.12 |  | 2.11 |  |  | 0.26 |  |
| Village volunteer | 0.22 |  | 22.84 | 64.41 |  | 4.99 | 14.06 |
| Demonstration participant | 0.22 |  |  | 13.97 |  |  | 3.05 |
| PIC impregnator | 0.22 |  |  | 69.87 |  |  | 15.25 |
| Neighborhood helper | 0.22 |  |  | 174.68 |  |  | 38.14 |
| Other village residents attending demonstration | 0.14 |  |  | 27.29 |  |  | 3.82 |
| Tailor | 0.68 |  |  | 3.49 |  |  | 2.36 |
| Tailor's helper | 0.23 |  |  | 3.49 |  |  | 0.79 |
| Sprayer | 1.53 |  |  | 9.36 |  |  | 14.30 |
| *Subtotal personnel* |  |  |  |  | 66.6 | 17.2 | 167.4 |
| *Non-personnel* |  |  |  |  |  |  |  |
| Expanded beads (kg) | 3.8 |  | 28.2 |  |  | 107.8 |  |
| Curtain fabric (m<sup>2</sup>) | 0.49 |  |  | 351.6 |  |  | 173.7 |
| Other for EPB treatment | 0.004 |  | 1100 |  |  | 4.4 |  |
| Other for PIC treatment | 0.020 |  |  | 1025 |  |  | 20.5 |
| *Subtotal non-personnel* |  |  |  |  | 0.00 | 112.2 | 194.1 |
| *Total (all resources)* |  |  |  |  | **66.6** | **129.5** | **361.5** |

Notation: MDA denotes mass drug administration; EPB denotes expanded polystyrene beads; PIC denotes pyrethroid impregnated curtains; VC denotes vector control; hr denotes hour; kg denotes kilograms, m<sup>2</sup> denotes square meter. Note: Total costs equal unit cost times the quantity. Quantities shown in the table are rounded.

**Table 4:** Annualized costs (US$) per 1,000 population at the block primary health care level.

| Input | Unit cost | Number of units | | | Total cost (US\$) | | |
| --- | --- | --- | --- | --- | --- | --- | --- |
|  |  | MDA | EPB | PIC | MDA | EPB | PIC |
| <u>Block primary health center staff (hours)</u> |  |  |  |  |  |  |  |
| Medical officers | \$4.8 | 2.3 | 0.4 | 0.4 | 11.3 | 2.1 | 2.1 |
| Public health supervisors | \$4.5 | 3.2 | 0.7 | 0.7 | 14.4 | 3.0 | 3.0 |
| Nurses | \$4.0 | 2.6 | | | 10.2 | | |
| Health inspectors | \$3.5 | 4.7 | 3.0 | 3.0 | 16.5 | 10.7 | 10.7 |
| Assistant field workers | \$2.5 | 1.7 | | | 4.1 | | |
| Administration workers | \$2.5 | 1.2 | | | 2.9 | | |
| Press people | \$2.2 | 0.5 | | | 1.1 | | |
| Personnel others | \$5.3 | 7.7 | | | 41.2 | | |
| <u>VC project staff (hours)</u> |  |  |  |  |  |  |  |
| Lead persons | \$5.5 | | 5.8 | 5.8 | | 31.9 | 31.9 |
| Project officers | \$1.6 | | 11.4 | 11.4 | | 17.6 | 17.6 |
| Technicians | \$0.6 | | 15.0 | 15.0 | | 9.6 | 9.6 |
| Project assistants | \$0.4 | | 90.9 | 90.9 | | 31.8 | 31.8 |
| Project administrators | \$1.1 | | 3.8 | 3.8 | | 4.3 | 4.3 |
| Volunteers | \$0.01 | | 930.0 | 930.0 | | 9.3 | 9.3 |
| *Subtotal Personnel* |  |  |  |  | 101.7 | 120.2 | 120.2 |
| <u>Non-personnel</u> |  |  |  |  |  |  |  |
| Expanded beads (kg) | \$0.1 | | 1010.0 | | | 111.1 | |
| Curtain fabric (square meter) | \$0.2 | | | 998.9 | | | 189.8 |
| Non-personnel, other | \$1.0 | | 108.6 | 110.1 | | 108.6 | 110.1 |
| *Subtotal Non personnel* |  |  |  |  | 0.0 | 219.6 | 299.8 |
| *Total (all resources)* |  |  |  |  | **101.7** | **339.8** | **420.0** |

Notation: MDA denotes mass drug administration; EPB denotes expanded polystyrene beads; PIC denotes pyrethroid impregnated curtains; VC denotes vector control; hr denotes hour; kg denotes kilograms, n.a. denotes not applicable (various inputs). Notes: Total costs equals unit cost times quantity. Volunteers were not paid for vector control activities, but occasionally received refreshments. Unit costs for personnel are hourly payments for paid staff, and average hourly refreshment costs for volunteers. Non-personnel other is a composite of all other inputs, including transportation. The quantities shown are rounded.

**Table 5:** Economic cost of interventions (US$) by funder and management level (<u>1,000 population)</u>.

| Management level | <u>MDA</u> |  | <u>EPB</u> |  | <u>PIC</u> |  |
| --- | --- | --- | --- | --- | --- | --- |
|  | Gov't | Comm. | Gov't | Comm. | Gov't | Comm. |
| 1. State | 208.66 | 0.00 | 0.02 | 0.00 | 0.02 | 0.00 |
| 2. HUD | 3.58 | 0.00 | 1.29 | 0.00 | 1.67 | 0.00 |
| 3. Block | 101.69 | 0.00 | 339.82 | 0.00 | 420.01 | 0.00 |
| 4. Subcenter | 99.24 | 0.00 | 20.34 | 0.00 | 20.34 | 0.00 |
| 5. Village | 0.14 | 66.58 | 0.00 | 129.46 | 0.00 | 361.47 |
| 6. 50 households | <u>9.30</u> | <u>44.36</u> | <u>0.00</u> | <u>0.00</u> | <u>0.00</u> | <u>0.00</u> |
| Subtotal | 422.61 | 110.94 | 361.46 | 129.46 | 442.04 | 361.47 |
| Combined total |  | 533.55 |  | 490.92 |  | 803.51 |

Notation: MDA denotes mass drug administration; EPB denotes expanded polystyrene beads; PIC denotes pyrethroid impregnated curtains; Gov't denotes government; Comm. denotes community; HUD denotes health unit district. Combined total sums costs funded by government and communities.

### Proportions of inputs and activities

Figure 2 presents the breakdown of costs by activity and intervention. Among the three interventions, PIC is the costliest. Among activities within each intervention, procurement of the materials and delivery of the service (termed procurement and delivery) was the costliest category. The non-personnel costs of VC were much higher than those of the MDA program. As EPB and PIC are supplements to MDA, the arms with these interventions cost more than MDA alone.

**Figure 2:**
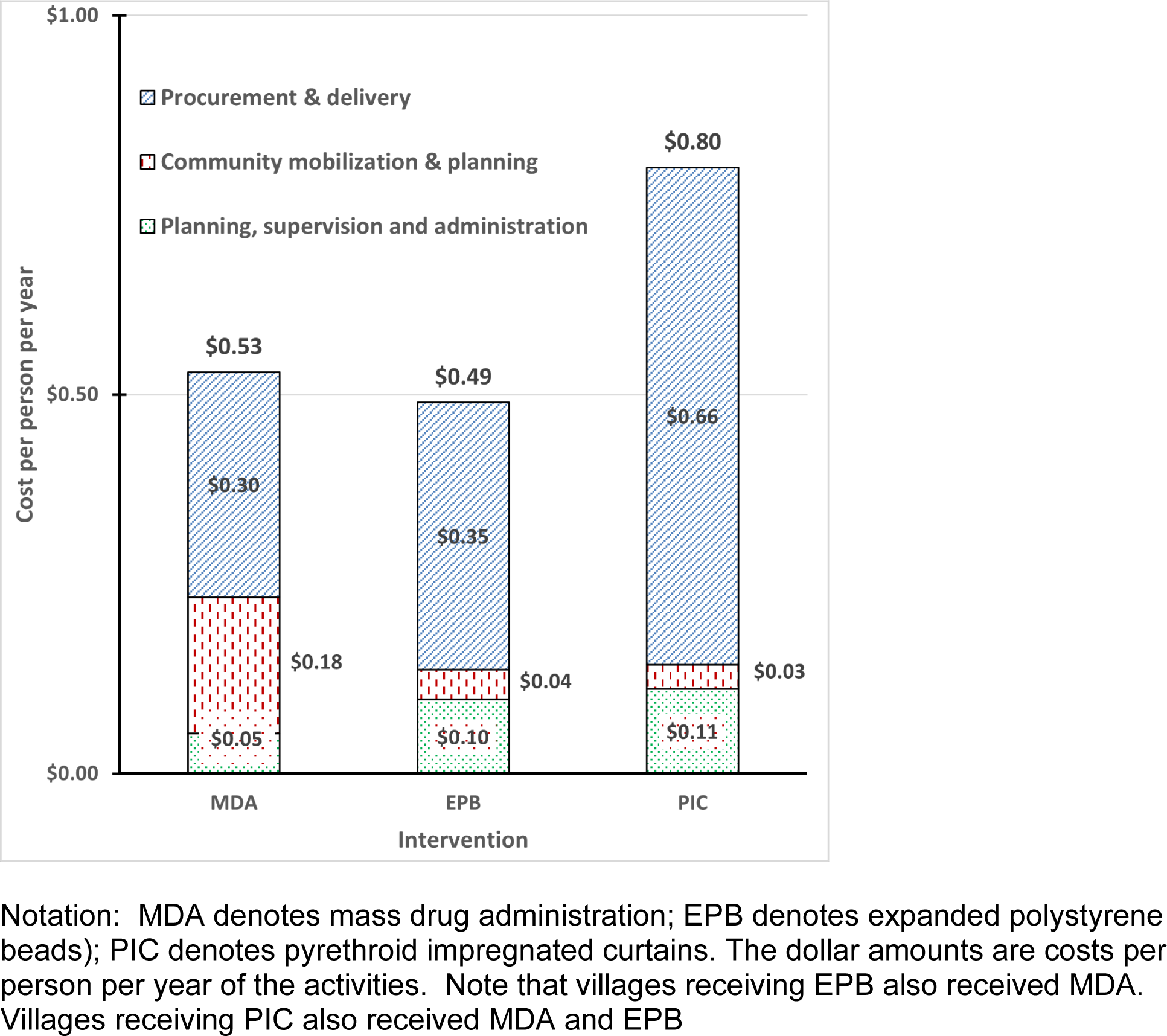
Cost per person per year by intervention and activity (US$)

### Progress towards LF elimination in the study area

Descriptive analyses showed a decline in microfilaria prevalence rates (MfP) for village groups within each of the treatment conditions from 2010 through the end of CRME‘s collection of entomological data in February 2013. Under MDA-alone, MfP fell from its baseline levels (± standard error of the mean, using the approximation of a normally distributed variable) by 84 (±25) percentage points. Where MDA was augmented by VC, villages with VCS reduced MfP by 75 (±20) and villages with VCI reduced MfP by 90 (±22) percentage points, respectively. All reductions compared to baseline levels were statistically significant [27]. Similarly, the “disease-free” share of population (i.e., living in villages with MfP below 1%) increased from 17% to 83% (66 percentage point improvement) in villages with MDA alone, from 8% to 67% (59 percentage point improvement) in VCS villages, and from 17% to 92% in VCI villages (75 percentage point improvement).

The percentages of villages with elimination are shown in Figure 3 for MfP and Figure 4 for AgP. Figure 5 shows the average AgP by study area and year. Similar to the results for average prevalence, all arms showed substantial improvement over time. However, the ranking of arms varied by year and by indicator, suggesting that no single arm had dramatically superior results.

**Figure 3:**
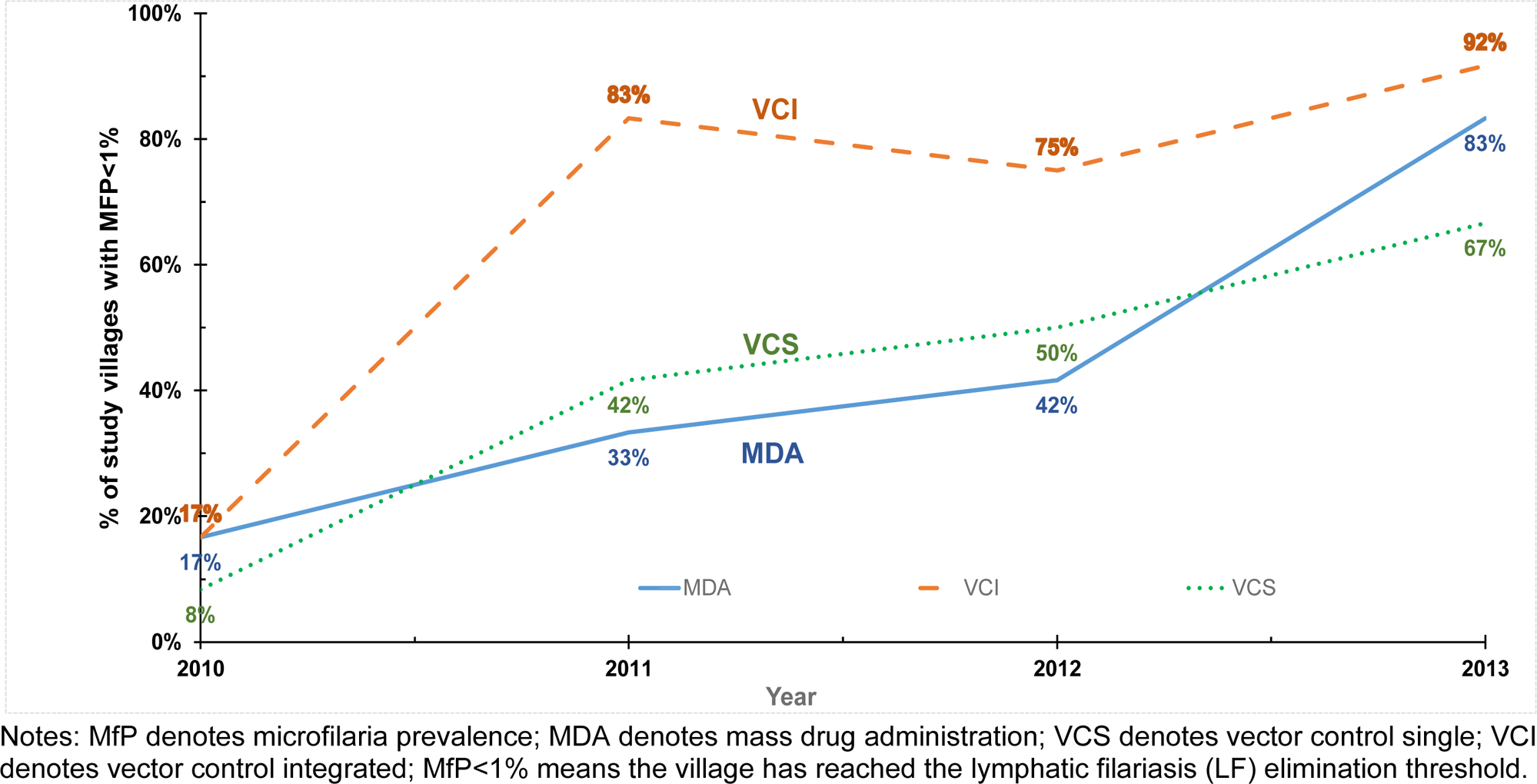
Share of villages achieving micro-filaria prevalence (MfP) target by intervention arm

**Figure 4:**
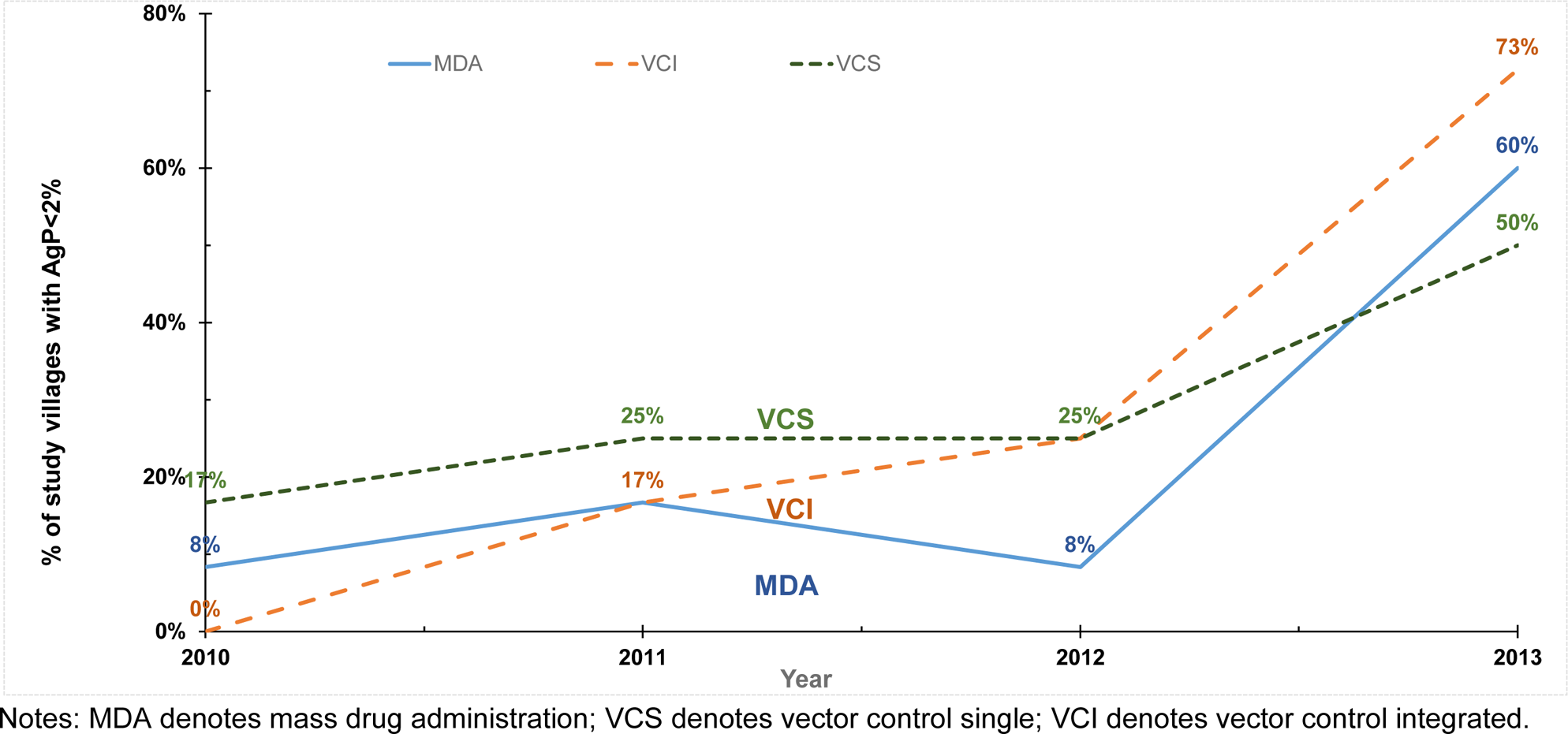
Share of villages achieving antigen prevalence (AgP) target by intervention arm

**Figure 5:**
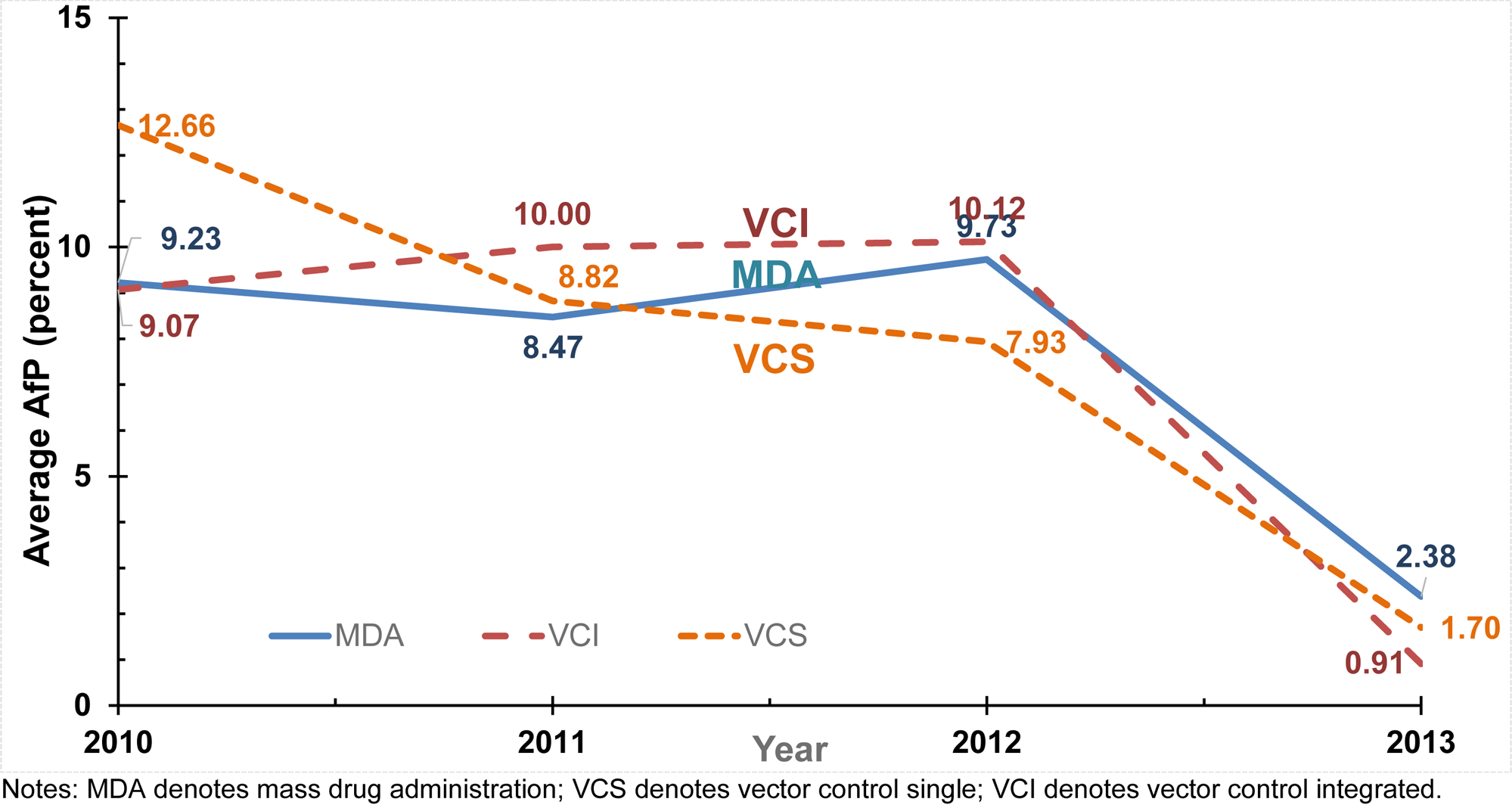
Average antigen prevalence (AgP) by intervention arm

Using the AgP measure for each intervention, the relative reductions from 2010 to 2013 were similar to those for MfP: 70 percentage points for MDA-alone, 89 percentage points for VCS, and 79 percentage points for VCI.

### Elimination results and context

Table 6 summarizes the results of the logistic regressions on the effectiveness of the two LF measures. The variable Year2013 is highly significant, indicating marked progress towards elimination in all arms. However, none of the coefficients for interactions between Year2013 and intervention were statistically significant. Furthermore, for each VC arm (VCS and VCI), the coefficients for MfP and AgP were of opposite sign. This lack of consistency and absence of statistical significance indicates that the differences between MDA alone and MDA supplemented by VC initiatives were consistent with chance. That is, at the time and place of the randomized trial, neither VCS nor VCI offered any statistically significant incremental benefit.

**Table 6:**
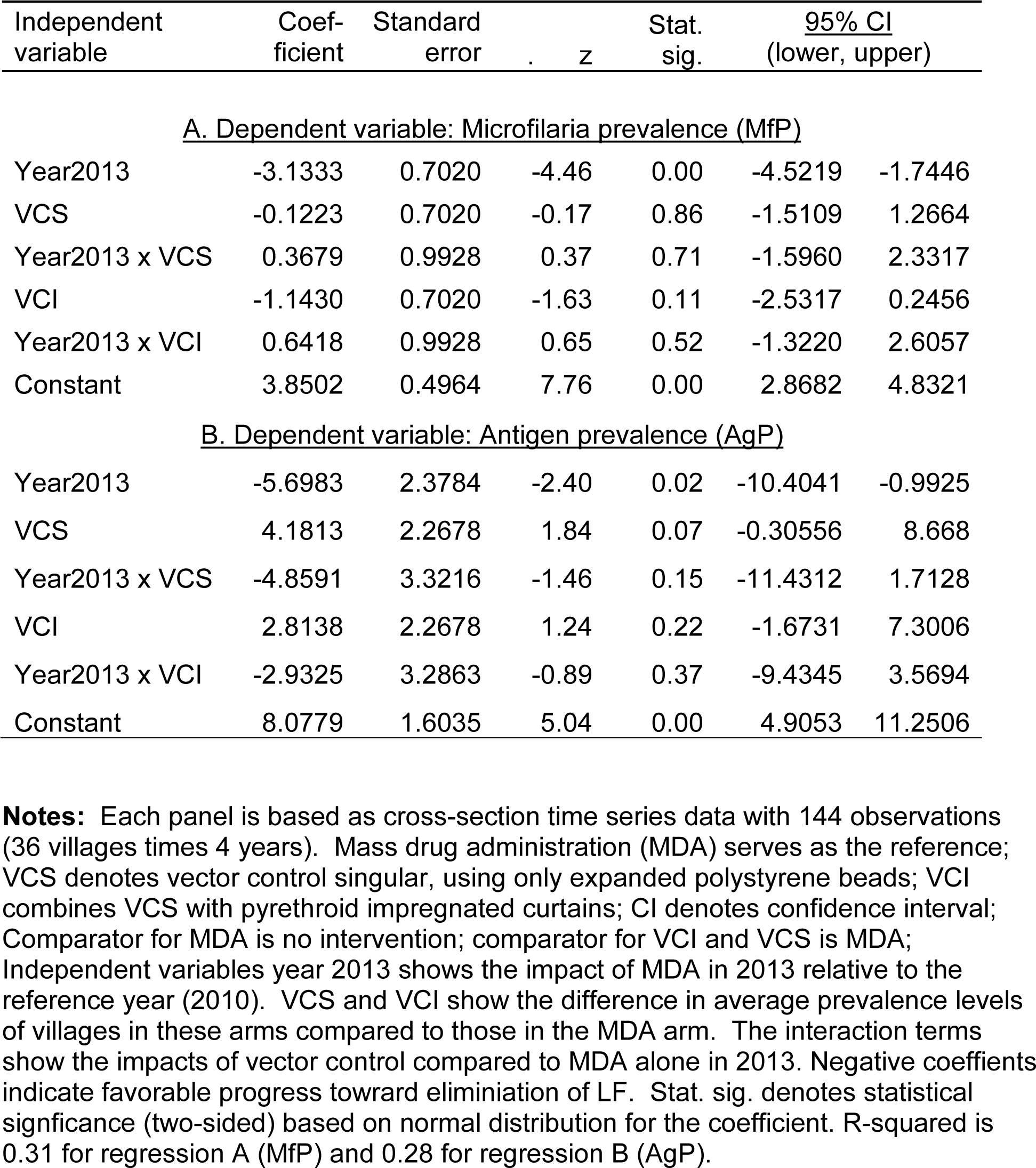
Linear regression results of incremental impact of vector control.

| Independent variable | Coef-<br>ficient | Standard<br>error | z | Stat.<br>sig. | <u>95% CI</u><br>(lower, upper) |  |
| --- | --- | --- | --- | --- | --- | --- |
| <u>A. Dependent variable: Microfilaria prevalence (MfP)</u> |  |  |  |  |  |  |
| Year2013 | -3.1333 | 0.7020 | -4.46 | 0.00 | -4.5219 | -1.7446 |
| VCS | -0.1223 | 0.7020 | -0.17 | 0.86 | -1.5109 | 1.2664 |
| Year2013 x VCS | 0.3679 | 0.9928 | 0.37 | 0.71 | -1.5960 | 2.3317 |
| VCI | -1.1430 | 0.7020 | -1.63 | 0.11 | -2.5317 | 0.2456 |
| Year2013 x VCI | 0.6418 | 0.9928 | 0.65 | 0.52 | -1.3220 | 2.6057 |
| Constant | 3.8502 | 0.4964 | 7.76 | 0.00 | 2.8682 | 4.8321 |
| <u>B. Dependent variable: Antigen prevalence (AgP)</u> |  |  |  |  |  |  |
| Year2013 | -5.6983 | 2.3784 | -2.40 | 0.02 | -10.4041 | -0.9925 |
| VCS | 4.1813 | 2.2678 | 1.84 | 0.07 | -0.30556 | 8.668 |
| Year2013 x VCS | -4.8591 | 3.3216 | -1.46 | 0.15 | -11.4312 | 1.7128 |
| VCI | 2.8138 | 2.2678 | 1.24 | 0.22 | -1.6731 | 7.3006 |
| Year2013 x VCI | -2.9325 | 3.2863 | -0.89 | 0.37 | -9.4345 | 3.5694 |
| Constant | 8.0779 | 1.6035 | 5.04 | 0.00 | 4.9053 | 11.2506 |

**Notes:** Each panel is based as cross-section time series data with 144 observations (36 villages times 4 years). Mass drug administration (MDA) serves as the reference; VCS denotes vector control singular, using only expanded polystyrene beads; VCI combines VCS with pyrethroid impregnated curtains; CI denotes confidence interval; Comparator for MDA is no intervention; comparator for VCI and VCS is MDA; Independent variables year 2013 shows the impact of MDA in 2013 relative to the reference year (2010). VCS and VCI show the difference in average prevalence levels of villages in these arms compared to those in the MDA arm. The interaction terms show the impacts of vector control compared to MDA alone in 2013. Negative coefficients indicate favorable progress toward elimination of LF. Stat. sig. denotes statistical significance (two-sided) based on normal distribution for the coefficient. R-squared is 0.31 for regression A (MfP) and 0.28 for regression B (AgP).

As context, Figure 6 presents both the global and India national prevalence of LF from 1990 through 2019 based on the GBD study [52]. Over this period of almost three decades, the prevalence of LF declined by three quarters both globally and in India. Yet the prevalence rate in India remained about triple the global average. The latest (2019) prevalence rates of 2.7% in India and 0.9% globally translate to 37.2 and 71.9 million people affected in India and globally respectively. Of all people affected globally, 52% are in India, indicating that LF remained a serious public health problem in India. By dividing the DALY burden by the prevalence of LF in the GBD study, we found that each person-year of LF carried a 0.022 DALY burden. In other words, every 45 persons with LF constitute the loss in value equivalent to one year of good health (i.e., 1/0.022 = 45).

**Figure 6.**
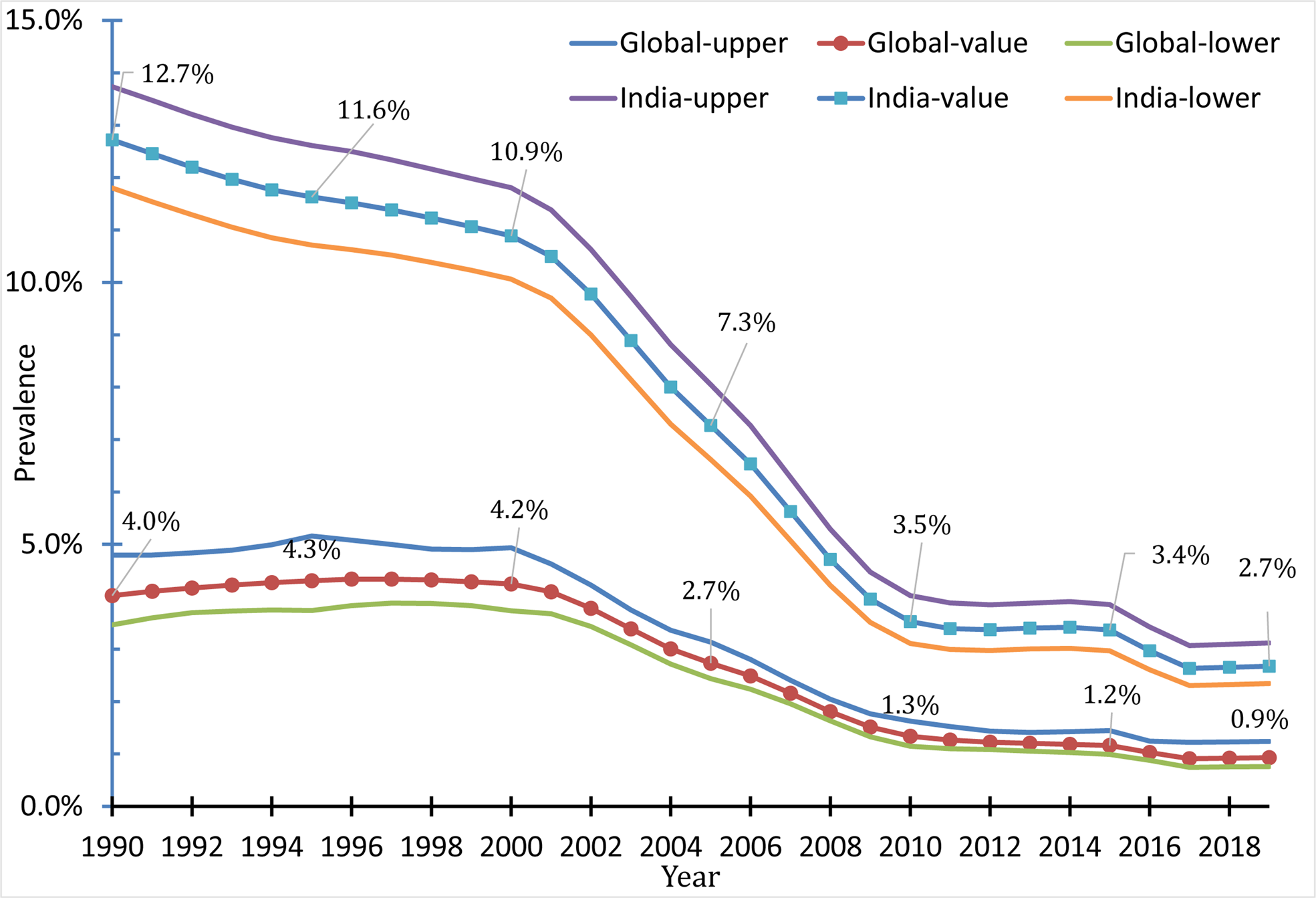
Prevalence of lymphatic filariasis by year globally and in India, 1990-2019

### Incremental cost-effectiveness ratio and summary results

Table 7 derives the ICER of MDA, the one intervention that was significantly effective. MDA significantly improved (lowered) both MfP and AfP compared to initial values 3 years earlier. The ICER for MDA is $112 per DALY. This ICER is substantially lower than India’s 2010 threshold of $1,350.6 per capita GDP in 2010 US dollars from the World Bank [53].

**Table 7.** Derivation of the incremental cost-effectiveness ratio of MDA (2010 US dollars)

| Row | Component | Source | Value |
| --- | --- | --- | --- |
| (1) | Baseline (2010) % of villages with MfP<1% | Figure 3 | 17% |
| (2) | Final (2013) % of villages with MfP<1% | Figure 3 | 83% |
| (3) | Cumulative change in % of villages with MfP<1% | (2) - (1) | 66% |
| (4) | Average annual increase in % of villages with MfP<1% | (3) / 3 | 22.0% |
| (5) | Baseline (2010) % of villages with AgP<2% | Figure 4 | 8% |
| (6) | Final (2013) % of villages with AgP<2% | Figure 4 | 60% |
| (7) | Cumulative change in % of villages with AfP<2% | (6) - (5) | 52% |
| (8) | Average annual increase in % of villages with AfP<2% | (7) / 3 | 17.3% |
| (9) | Pooled annual increase in % of villages with MfP<1% and AfP<2% | [(4) + (8)] / 2 | 19.7% |
| (10) | Implicit comparator to MDA | Text | No prevention |
| (11) | Incremental % of villages with MfP<1% and AgP<2% | (9) - (10) | 19.7% |
| (12) | DALYs per person with lymphatic filariasis (LF) | GBD Study | 0.022 |
| (13) | DALYs per 1,000 persons with LF | 1,000 x (12) | 22 |
| (14) | DALYs averted per 1,000 persons in arm | (11) x (13) | 4.33 |
| (15) | Overall cost per 1,000 population | Table 5 | \$533.35 |
| (16) | Incremental cost per 1,000 population | (15) - (10) | \$533.35 |
| (17) | Incremental cost-effectiveness ratio (ICER), \$/DALY | (16) / (14) | \$112 |
| (18) | India's 2010 per capita Gross Domestic Product (GDP) in 2010 US\$ | World Bank† | \$1,350.6 |
| (19) | ICER as fraction of India's per capita GDP | (17) / (18) | 8% |

Notes: DALY denotes disability adjusted life years; MDA denotes mass drug administration; MfP denotes microfilaria prevalence in village during the year; AgP denotes antigen prevalence in the village during the year. † Source: The World Bank[53].

## Discussion

This study documents substantial progress in controlling LF in the study area from 2010 through 2013, as well as over a period of almost 3 decades across all of India and globally. This progress was achieved largely through MDA. Yet, despite MDA, an estimated 37.2 million people in India remained affected by LF in the latest data (2019) [52]. A key policy question is whether existing control strategies are sufficient, or whether VC must be added as well.

This study was planned when the impact of MDA on LF had slowed, and MDA appeared insufficient to be able to eliminate LF as a public health problem in India. Supplementing MDA by one or both VC strategies was hypothesized to make a potentially crucial difference. The goals of this study were to assess the cost, effectiveness, and cost-effectiveness of MDA and the VC strategies as complements to MDA.

Prior to the study period, the impact of MDA was mixed. Tamil Nadu began single dose DEC mass drug administration (MDA) from 1997-98 in all endemic districts and switched to a two-drug regimen of DEC and albendazole in 2001 [54]. However, gaps in annual rounds occurred 1997, 2005 and 2006 in Tamil Nadu. Over the study period, however, MDA proved substantially more efficacious than it had previously. We counted 11 completed MDA rounds for our study villages in Villupuram District during the 15-year period ending in 2012. According to WHO’s protocols for LF chemotherapy [55], after five rounds of MDA the implementation units could stop the interventions if they had achieved ≥ 65% program coverage during each of the five rounds and had reached the target levels of MfP<1% or AgP<2%. A period of post-treatment surveillance would follow to be sure that LF transmission had been interrupted. In 2012, the district ended its MDA program [56].

The study data found that MDA was effective and highly cost-effective in controlling LF and progressing towards elimination. Improvements in adherence may explain the success of MDA in the study site compared to earlier expectations. An evaluation of Tamil Nadu’s MDA program in 2000 found mediocre adherence, with 30% households being missed and 46% of those covered not swallowing the recommended MDA tablets [50]. Our interviewees reported, however, that MDA delivery rates and compliance have progressively improved over the subsequent years, consistent with the steady declines in MfP and AgP under MDA alone. As MDA is delivered by community-level workers with support from higher levels in the health system, its efficacy demonstrates success of Tamil Nadu’s system of social mobilization and human development. The finding is consistent with the State’s ranking as one of India’s best on this dimension [57].

As far as we know, this study is the only comparative cost-effectiveness study of alternative LF control strategies. This study did not find a statistically significant incremental progress towards LF elimination as a public health problem from either VC intervention. We concluded that VCS and VCI were not significantly effective nor cost-effective additions to MDA during the study period in this setting. Thus, this study did not find quantitative evidence within the 3-year study period to support the addition of VC. Continuing with high coverage and full compliance with the regimen of medicines (MDA), rather than adding VC, proved to be the most effective use of limited resources.

The reason why VC failed to demonstrate incremental impact might be attributable to the presence of numerous untreated breeding sites for *Culex quinquefasciatus*, such as blocked drains, buckets, discarded containers and obscure disused cesspits, which yielded sufficient mosquitoes for continuing LF transmission. To address this challenge the NVBDCP has relied on more integrated VC strategies similar to VCI [58] but we are not aware of any cost-effectiveness evaluation.

After 2015, the Transmission Assessment Survey found that 222 of the 256 districts previously classified as endemic for LF had MfP adequately controlled [59]. Most of these districts have stopped their MDA programs as they were no longer needed. However, 12 such villages in non-endemic areas had MfP between 3.2% and 11.2% [60].

Qualitative observations and entomological data identified some potential benefits of VCI. Mosquito nuisance was reduced. Also, curtains can protect households from other insects in addition to mosquitoes, thereby reducing transmission risks for malaria, arboviruses, leishmaniasis and other infections. As VC is less dependent on individual behavior than MDA, it could potentially achieve higher coverage and compliance. Both VC interventions were reported to heighten the public’s awareness of LF and capacity to control it, and perhaps improved compliance with MDA [27]. However, unless these added benefits were quantified and proved to be substantial, they cannot justify the incremental expense of VC.

Two opposing study limitations must be acknowledged. On the one hand, our collection of data for costs of MDA was retrospective and may have missed hidden costs of some extended activities, such as more days for MDA distribution, and contributed time from other government departments. On the other hand, costs in this trial may have been higher than those that would have been routinely found. Donor funding allowed engaging more and higher skilled staff. As a first-time effort, some experimentation required additional time. If VC had been funded entirely from Indian sources, procurement procedures might have been faster and less expensive.

## Conclusion

Before-and-after data in this trial indicate that MDA was highly effective in controlling LF as measured by both MfP and AgP. This success left little scope for further improvement. With substantially reduced levels of LF, the study did not find any significant additional benefits that VC added to MDA over the 3-year study period. This study did not assess whether VC could have replaced MDA or proven more impactful over a longer timeframe. However, our cost data show that VC would probably remain more expensive than MDA. As India’s 2.7% LF prevalence persists at three times the global average [52], the goal of elimination justifies continuing MDA, possibly using triple-drug therapy [61], in communities where LF rates remain above 2% AgP or 1% MfP thresholds.

## Data Availability

All data produced in the present study are available upon reasonable request to the authors or in the authors' prior publications.

## Acknowledgments

We thank: Eric Ottesen, PJ Hooper, the late Dominique Kyelem and other personnel of the Task Force for Global Health for thoughtful ideas and logistical and financial support; the late S. Elango, S. Sreedharan, and M. Alamelu of the State Public Health Department of Tamil Nadu; M. Geetha, C. Palanichamy, and other health staff of the health unit district of Kallakurichi; Muthu Kumar, T. Sekar, T.A. Srinivasan, and other staff of the Ariyur Block PHC for providing key program and financial information; health inspectors, village health nurses, other medical officers, and village presidents, the late R. Rajendran, A. Munirathinam, S. Ravi, and V. Ashok Kumar of CRME for providing essential program data during our field visits; the volunteer program workers for their critical contributions to the MDA and VC services underlying this study; and Clare L. Hurley of Brandeis University for editorial support.

## Emails

Donald S Shepard, Ph.D.

Aung K Lwin M.D., M.S

Sunish I. Pulikkottil, Ph.D.

Mariapillai Kalimuthu, MS

Natarajan Arunachalam, Ph.D.

Brij K. Tyagi Ph.D.

Graham B. White Ph.D.

## Annex. Acronyms

AgP: Antigen prevalence
BLPHC: Block level PHC
CRME: Centre for Research in Medical Entomology
DALY: disability adjusted life year
DEC: diethylcarbamazine
ELF: elimination of lymphatic filariasis
EPB: expanded polystyrene beads
EPS: expanded polystyrene
FL: Florida (state, USA)
HUD: Health unit district
ICMR: Indian Council for Medical Research
ICT: immunochromatography card test
IRR: incident rate ratio
IVM: Integrated vector management
LF: Lymphatic filariasis
M: mean
MA: Massachusetts state, USA
M.D.: Doctor of Medicine
MDA: mass drug administration
MDA-CAT: MDA cost analysis tool
MfP: microfilaria prevalence
M.S.: Master of Science
NTD: neglected tropical disease
NVBDCP: National Vector Borne Disease Control Program
PHC: primary health center
Ph.D.: Doctor of Philosophy
PIC: pyrethroid impregnated curtain
SC: suspension concentrate
SD: standard deviation
SHD: State health department
TAS: transmission assessment survey
TN: Tamil Nadu State, India
TX: Texas State, USA
US$: United States dollar
USA: United States of America
USAID: United States Agency for International Development
VC: vector control
VC-CAT: MDA+ cost analysis tool
VCI: Vector control integrated
VCS: Vector control single
WHO: World Health Organization

## Supporting information legends

S1 Figure. Discussion of study plans at the Field Unit. From left to right: Prof. Donald S. Shepard, Dr B.K. Tyagi and Prof. Graham B. White

S2. Figure. Visit to a study village. From left to right: Shri R. Krishnamoorth (CRME Field Unit), Prof. Donald S. Shepard, Dr. Sunish Pulikkottil, Dr. Aung K. Lwin, Shri A. Munirathinam (CRME Field Unit)

## References

1. World Health Organization (WHO). Integrating neglected tropical diseases into global health and development: Fourth WHO report on neglected tropical diseases. Geneva: WHO; 2017. Available from: WHO/HTM/NTD/2017.01, 278pp.

2. Dhariwal AC, Srivastava PK, Bhattacharjee J. Elimination of lymphatic filariasis in India: An update. J Ind Med Assoc. 2015;113(12):189–90 PubMed Central PMCID: 19552103.

3. Lobo DA, Velayudhan R, Chatterjee P, Kohli H, Hotez PJ. The neglected tropical diseases of India and South Asia: Review of their prevalence, distribution, and control or elimination. PLoS Negl Trop Dis. 2011;5(10): e1222. doi:10.1371/journal.pntd.0001222.

4. Ramaiah KD, Das PK. Mass drug administration to eliminate lymphatic filariasis in India. Trends Parasitol. 2004;20(11):499–502. doi: 10.1016/j.pt.2004.08.010.

5. Tyagi BK (Ed.). Lymphatic Filariasis Epidemiology, Treatment and Prevention - The Indian Perspective. Singapore: Springer Nature; 2018. xix+333pp. https://link.springer.com/book/10.1007/978-981-13-391-2 p.

6. Kumar P, Ahmad S, Bhar D, Roy R, Singh B. ’Whenever I tell her to wear slippers, she turns a deaf ear. She never listens’: A qualitative descriptive research on the barriers to basic lymphedema management and quality of life in lymphatic filariasis patients in a rural block of eastern India Parasit Vectors. 2023;16(1):429. doi: 10.1186/s13071-023-06036-0.

7. World Health Organization. Global programme to eliminate lymphatic filariasis: progress report, 2021. Wkly Epidemiol Rec, 97, 513–524. 2022. Available from: https://iris.who.int/bitstream/handle/10665/363513/WER9741-eng-fre.pdf?sequence=.

8. World Health Organization (WHO). Lymphatic filariasis: Key facts. 2023. Available from: https://www.who.int/news-room/fact-sheets/detail/lymphatic-filariasis.

9. Agrawal VK, Sashindran VK. Lymphatic filariasis in India: problems, challenges and new initiatives. Med J Armed Forces India. 2006;62(4):359–62. doi: 10.1016/S0377-1237(06)80109-7.

10. Global Alliance to Eliminate Lymphatic Filariasis (GAELF). Global Alliance to Eliminate Lymphatic Filariasis, Report of the Second Meeting. New Delhi, India: 2-3 May 2002.

11. World Health Organization (WHO). Annual Report on Lymphatic Filariasis 2001. Global Programme to Eliminate Lymphatic Filariasis. 2002. Available from: WHO/CDS/CPE/CEE/2002.28, 81.

12. Kalimuthu M, Sunish IP, Nagaraj J, Munirathinam A, Ashok Kumar V, Arunachalam N, et al. Residual microfilaraemia in rural pockets of south India after five rounds of DEC plus albendazole administration as part of the LF elimination campaign. J Vector Borne Dis. 2015;52(6):182–4. PMID: 26119554.

13. Babu BV, Babu GR. Coverage of, and compliance with, mass drug administration under the programme to eliminate lymphatic filariasis in India: A systematic review. Trans R Soc Trop Med Hyg. 2014;108(9):538–49. doi: 10.1093/trstmh/tru057.

14. Jambulingam P, Subramanian S, de Vlas SJ, Vinubala C, Stolk WA. Mathematical modelling of lymphatic filariasis elimination programmes in India: required duration of mass drug administration and post-treatment level of infection indicators. Parasit Vectors 2016;9:501. doi 10.1186/s13071-016-1768-y.

15. Ramaiah KD, Das PK, Michael E, Guyatt H. The economic burden of lymphatic filariasis in India. Parasitol Today. 2000;16(6):251–3. doi: 10.1016/s0169-4758(00)01643-4.

16. Zagaria N, Savioli L. Elimination of lymphatic filariasis: A public-health challenge. Ann Trop Med Parasitol. 2002;96(Suppl 2):S3–13. doi: 10.1179/00034980215002347.

17. Raju K, Jambulingam P, Sabesan S, Vanamail P. Lymphatic filariasis in India: Epidemiology and control measures. J Postgrad Med. 2010;56(3):232–8. doi: 10.4103/0022-3859.68650.

18. World Health Organizaiton (WHO). Integrated vector management to control malaria and lymphatic filariasis: WHO position statement. 2011. Available from: https://apps.who.int/iris/handle/10665/70817.

19. World Health Organizaiton (WHO). Lymphatic filariasis: A handbook of practical entomology for national lymphatic filariasis elimination programmes. 2013. Available from: WHO/HTM/NTD/PCT/2013.10.

20. Bockarie MJ, Pedersen EM, White GB, Michael E. Role of vector control in the Global Program to Eliminate Lymphatic Filariasis. Annu Rev Entomol. 2009;54:469–87. doi: 10.1146/annurev.ento.54.110807.090626.

21. Ottesen EA, Duke BO, Karam M, Behbehani K. Strategies and tools for the control/elimination of lymphatic filariasis. Bull World Health Organ. 1997;75(6):491–503, PMID: 9509621.

22. Reuben R, Rajendran R, Sunish IP, Mani TR, Tewari SC, Hiriyan J, et al. Annual single dose diethylcarbamazine plus ivermectin for control of bancroftian filariasis: Comparative efficacy with and without vector control. Ann Trop Med Parasitol. 2007;95(4):361–78. doi: 10.1080/00034980120065796.

23. Sunish IP, Rajendran R, Mani TR, Munirathinam A, Tewari SC, Hiriyan J, et al. Resurgence in filarial transmission after withdrawal of mass drug administration and the relationship between antigenaemia and microfilaraemia - A longitudinal study. Trop Med Int Health. 2002;7(1): 59–69. doi: 10.1046/j.1365-3156.2002.00828.x.

24. Sunish IP, Rajendran R, Mani TR, Munirathinam A, Tewari SC, Hiriyan J, et al. Transmission intensity index to monitor filariasis infection pressure in vectors for the evaluation of filariasis elimination programmes. Trop Med Int Health. 2003;8(9):812–9. doi: 10.1046/j.1365-3156.2003.01109.x.

25. Krishnamoorthy K, Rajendran R, Sunish I, Reuben R. Cost-effectiveness of the use of vector control and mass drug administration, separately or in combination, against lymphatic filariasis. Ann Trop Med Parasitol. 2002;96(Supplement 2):S77–90. doi: 10.1179/000349802125002428.

26. Sunish IP, Rajendran R, Arunachalam N, Munirathinan A, Tyagi BK, Elango S, et al. Assessment of the Added Impact of Vector Control for Augmentation of MDA to Eliminate Lymphatic Filariasis in South India. Final Report, August 2008 to July 2013. BMGF Funded project through RCC-ELF Grant 43922. Centre for Research in Medical Entomology (Indian Council for Medical Research), Madurai, Tamil Nadu. 2014.

27. Sunish I, Kalimuthu M, Ashok Kumar V, Munirathinam A, Nagaraj J, Tyagi BK, et al. Can community-based integrated vector control hasten the process of LF elimination? Parasitol Res. 2016;115:2353–62. doi: 10.1007/s00436-016-4985-6.

28. Sunish IP, Rajendran R, Mani TR, Munirathinam A, Dash AP, Tyagi BK. Vector control complements mass drug administration against Bancroftian filariasis in Tirukoilur, India. Bull World Health Organ. 2007;85:138–45. doi: 10.2471/blt.06.029389.

29. CCI. Census of India 2011, Provisional Population Total. Census Commissioner India, Office of the Registrar General, Ministry of Home Affairs, Government of India, New Delhi. 2011.

30. Sabesan S, Raju KH, Subramanian S, Srivastava PK, Jambulingam P. Lymphatic filariasis transmission risk map of India, based on a geo-environmental risk model. Vector Borne Zoonotic Dis. 2013;13(9):657–65. doi: 10.1089/vbz.2012.1238.

31. Subramanian NT. Annual Public Health Administration Report - 2008-09. Chennai: Government of Tamil Nadu, 2009.

32. National Health Mission Tamil Nadu. Department of Health and Family Welfare, State Health Society, Chennai. Available from: http://www.nrhmtn.gov.in/hud.html.

33. World Health Organizaiton (WHO). Monitoring and Epidemiological Assessment of Mass Drug Administration in the Global Programme to Eliminate Lymphatic Filariasis: A Manual for National Elimination Programmes. 2011. Available from: WHO/HTM/NTD/PCT/2011.4.

34. Rebollo M, Bockarie M. Toward the elimination of lymphatic filariasis by 2020: treatment update and impact assessment for the endgame. Expert Rev Anti Infect Ther. 2013;11(7):723–31. doi: 10.1586/14787210.2013.811841.

35. Rajendran R, Sunish IP, Munirathinam A, Kumar VA, Tyagi BK. Role of community empowerment in the elimination of lymphatic filariasis in south India. Trop Biomed. 2010;27(1):68–78. PMID: 20562816.

36. Bajpai N, Dholakia RH, Sachs JD. Scaling Up Primary Health Services in Rural Tamil Nadu: Public Investment Requirements and Health Sector Reform. New York: The Earth Institute at Columbia University, Center on Globalization and Sustainable Development Working Paper No.36, 65p. Academic Commons, 10.7916/D8R49XMW. 2008.

37. GlaxoSmithKline. Albendazole donation programme. Available from: https://www.gsk.com/media/3350/albendazole-donation-programme-infographic.pdf.

38. World Health Organizaiton (WHO). Control of Neglected Tropical Diseases. Global Programme to Eliminate Lymphatic Filariasis. 2023. Available from: https://www.who.int/teams/control-of-neglected-tropical-diseases/lymphatic-filariasis/global-programme-to-eliminate-lymphatic-filariasis.

39. Maxwell CA, Curtis CF, Haji H, Kisumku S, Thalib AI, Yahya SA. Control of Bancroftian filariasis by integrating therapy with vector control using polystyrene beads in wet pit latrines. Trans R Soc Trop Med Hyg. 1990;84(5):709–14. doi: 10.1016/0035-9203(90)90158-b.

40. Curtis C, Lines J, Carnevale P, Robert V, Boudin C, Halna J, et al. Impregnated bed nets and curtains against malaria mosquitoes. In: Curtis C, editor. Appropriate Technology in Vector Control. Boca Raton FL: CRC Press; 1990. pp. 5–46.

41. Curtis CF, Malecela-Lazaro M, Reuben R, Maxwell CA. Use of floating layers of polystyrene beads to control populations of the filaria vector Culex quinquefasciatus. Ann Trop Med Parasitol. 2002;96(Suppl 2):S97–S104. doi: 10.1179/000349802125002446.

42. Government of India. National Conter for Vector Borne Disease Control. 2023. Available from: https://ncvbdc.mohfw.gov.in/.

43. Frye JE. International Drug Price Indicator Guide, 2009 edition. Cambridge, MA: Management Sciences for Health, 2010.

44. Linehan M, Hanson C, Weaver A, Baker M, Kabore A, Zoerhoff KL, et al. Integrated implementation of programs targeting neglected tropical diseases through preventive chemotherapy: Proving the feasibility at national scale. Am J Trop Med Hyg. 2011;84(1):5–14. doi: 10.4269/ajtmh.2011.10-0411.

45. World Health Organizaiton (WHO). Neglected Tropical Diseases. 2017. Available from: http://www.who.int/neglected_diseases/diseases/en/.

46. XE. Current and Historical Rate Tables - 17 Jan 2010 - Indian Rupee. Available from: http://www.xe.com/currencytables/?from=INR&date=2010-01-17.

47. Mladonicky JM, King JD, Liang JL, Chambers E, Pa’au M, Schmaedick MA, et al. Assessing transmission of lymphatic filariasis using parasitologic, serologic, and entomologic tools after mass drug administration in American Samoa. Am J Trop Med Hyg. 2009;80(5):769–73. PMID: 19407122.

48. Pani SP, Hoti SL, Vanamail P, Das LK. Comparison of an immunochromatographic card test with night blood smear examination for detection of Wuchereria bancrofti microfilaria carriers. Natl Med J India. 2004;17(6):304–6. PMID: 15736550.

49. Schuetz A, Addiss DG, Eberhard ML, Lammie PJ. Evaluation of the whole blood filariasis ICT test for short-term monitoring after antifilarial treatment. Am J Trop Med Hyg. 2000;62(4):502–3. doi: 10.4269/ajtmh.2000.62.502.

50. Ramaiah KD, Das PK, Appavoo NC, Ramu K, Augustin DJ, Kumar KN, et al. A programme to eliminate lymphatic filariasis in Tamil Nadu state, India: compliance with annual single-dose DEC mass treatment and some related operational aspects. Trop Med Int Health. 2000;5(12):842–7. doi: 10.1046/j.1365-3156.2000.00659.x.

51. Turner HC, Bettis AA, Chu BK, McFarland DA, Hooper PJ, Ottesen EA, et al. The health and economic benefits of the global programme to eliminate lymphatic filariasis (2000–2014). Infect Dis Poverty 2016;5:54. doi: 10.1186/s40249-016-0147-4.

52. Institute of Health Metrics and Evaluation. Global Health Data Exchange. Lymphatic filariasis. 2023. Available from: https://vizhub.healthdata.org/gbd-results/.

53. The World Bank. GDP per capita (current US $) - India. 2023. Available from: https://data.worldbank.org/indicator/NY.GDP.PCAP.CD?locations=IN

54. Nandha B, Krishnamoorthy K, Jambulingam P. Towards elimination of lymphatic filariasis: Social mobilization issues and challenges in mass drug administration with anti-filarial drugs in Tamil Nadu, South India. Health Educ Res. 2013;28(4):591–8. doi: 10.1093/her/cyt042.

55. Chu BK, Deming M, Biritwum N-K, Bougma WR, Dorkenoo AM, El-Setouhy M, et al. Transmission assessment surveys (TAS) to define endpoints for lymphatic filariasis mass drug administration: A multicenter evaluation. PLoS Negl Trop Dis. 2013;203:e2584. doi:10.1371/journal.pntd.0002584.

56. National Health Mission Tamil Nadu, Department of Health and Family Welfare. Vector Borne Disease Control Programme, State Health Society, Chennai. Available from: http://www.nrhmtn.gov.in/vbdc.html.

57. Drèze J, Sen A. An Uncertain Glory: India and its Contradiction. Princeton University Press, NJ: Princeton University Press, NJ; 2013.

58. Srivastava PK, Sonal GS, Sharma SN, Singh S, Baruah K. Integrated Vector Management: Scenario Under National Vector Borne Disease Control Program, India. J Ind Med Assoc. 2015;113(12):179–82.

59. Srivastava PK, Dhariwal AC. Lymphatic Filariasis Elimination: Update for Mission Possible. 2018. Available from: https://link.springer.com/chapter/10.1007/978-981-13-1391-2_2.

60. Srividya A, Subramanian S, Jambulingam P, Vijayakumar B, Raja JD. Mapping and monitoring for a lymphatic filariasis elimination program: A systematic review. Res Rep Trop Med. 2019;10:43–90. doi: 10.2147/RRTM.S134186.

61. Tripathi B, Roy N, Dhingra N. Introduction of triple-drug therapy for accelerating lymphatic filariasis elimination in India: lessons learned. Am J Trop Med Hyg, 2022;106(Suppl 5): 29–38.

